# Inequities in hypertension management: an observational cross-sectional study in North-East London using electronic health records

**DOI:** 10.1101/2023.03.14.23286419

**Authors:** Stuart Rison, Oliver Redfern, Isabel Dostal, Chris Carvalho, Rohini Mathur, Zahra Raisi-Estabragh, John Robson

## Abstract

**Background:** Hypertension is a key potentially modifiable risk factor for cardiovascular disease-the leading cause of death in the UK. Good blood pressure (BP) control reduces mortality; however, health inequalities may lead to variability in hypertension management.

**Aim:** To investigate health inequities related to ethnicity, sex, age, and socio-economic status in the treatment and control of BP in a large cohort of adult patients with hypertension.

**Design and Setting:** A cross-sectional cohort study of adults with hypertension registered with general practices in North East London on 1^st^ April 2019.

**Method:** Multivariable logistic regression was used to estimate associations of demographics and treatment intensity on the following hypertension management indicators: 1) BP recording in last 12 months, 2) BP on age-adjusted target, 3) BP on age-adjusted target and BP recorded in last 12 months.

**Results:** In total, 156,296 adults were included. Black/Black British ethnic groups were less likely to have controlled BP than White ethnic groups (OR 0.89, 95% CI = 0.86-0.92). Asian/Asian British ethnic groups were more likely to have controlled blood pressure (OR 1.29, 95% CI = 1.25-1.34). Ethnic differences in control could not be explained by the likelihood of having a recent blood pressure recording, nor by treatment intensity differences. Older adults were more likely to have controlled hypertension than younger patients.

**Conclusion:** Black/Black British and younger people are less likely to have controlled hypertension and may warrant targeted interventions. Possible explanations for these findings are presented and further research is needed on reasons for ethnic differences.

**HOW THIS FITS IN:** Health inequities in the management of long-term conditions are widely recognised. This study identifies ethnic, age, gender and deprivation inequities in the control of blood pressure in a large unselected cohort of adults with hypertension in an ethnically diverse and nationally disadvantaged area of London. It confirms previous findings that blood pressure control in Black ethnic groups with hypertension is worse than in White or Asian ethnic groups. These differences were not related to access to blood pressure recording or treatment intensity. Younger adults were less likely to have controlled hypertension than older adults.

## INTRODUCTION

The importance of effective control of blood pressure in people with hypertension is recognised at local, national and international level as a major contributor to reduction in CVD.[1–4] Cardiovascular disease (CVD) remains the leading cause of death in the UK. In the last decade, the reduction in CVD mortality has stalled and for some disadvantaged groups, has actually increased.[5–7]

Health inequities in the management of hypertension have been investigated in multiple UK populations. In particular Black ethnic groups are significantly less likely to have controlled blood pressure than White ethnic groups and in some studies Asian patients had better control.[8–10] Similar inequities have been described in the USA and Europe.[11,12]

The localities in North East London in this study are among the most ethnically diverse and deprived in the United Kingdom.[13] However, supported by local quality improvement programmes, the control of blood pressure was the best in England in the national QOF rankings up until the pandemic.[14]

Our study included adults with hypertension and aimed to analyse social and demographic inequities in recording and control of blood pressure and treatment intensity.

## METHODS

A cross-sectional study was carried out in five North-East London Clinical Commissioning Groups (CCGs): City and Hackney (CH), Newham (NH), Redbridge (RB), Tower Hamlets (TH) and Waltham Forest (WF) of currently registered patients in general practices using the EMIS electronic health record system. Pseudonymised data were centrally extracted by the Clinical Effectiveness Group (CEG), Queen Mary University of London.

The study cohort comprised adults aged 18 years and older, with diagnosed hypertension on the index date, 1^st^ April 2019. The national NHS Quality and Outcomes Framework (QOF) codeset identified hypertension excluding “hypertension resolved” (Supplementary Table 1 and https://clinicalcodes.rss.mhs.man.ac.uk/medcodes/article/200/).

For each cohort individual, the following demographic data were extracted (Supplementary Table 2): Age in years on index date; Sex; home Lower Layer Super Output Areas (LSOA); Ethnic group code.

The most recent systolic and diastolic blood pressure values (mmHg) within 12 months of the index date and entry dates were extracted. Values were defined as missing for patients with incomplete, unreliable or unfeasible blood pressures (**Figure 1**).

**Figure 1:**
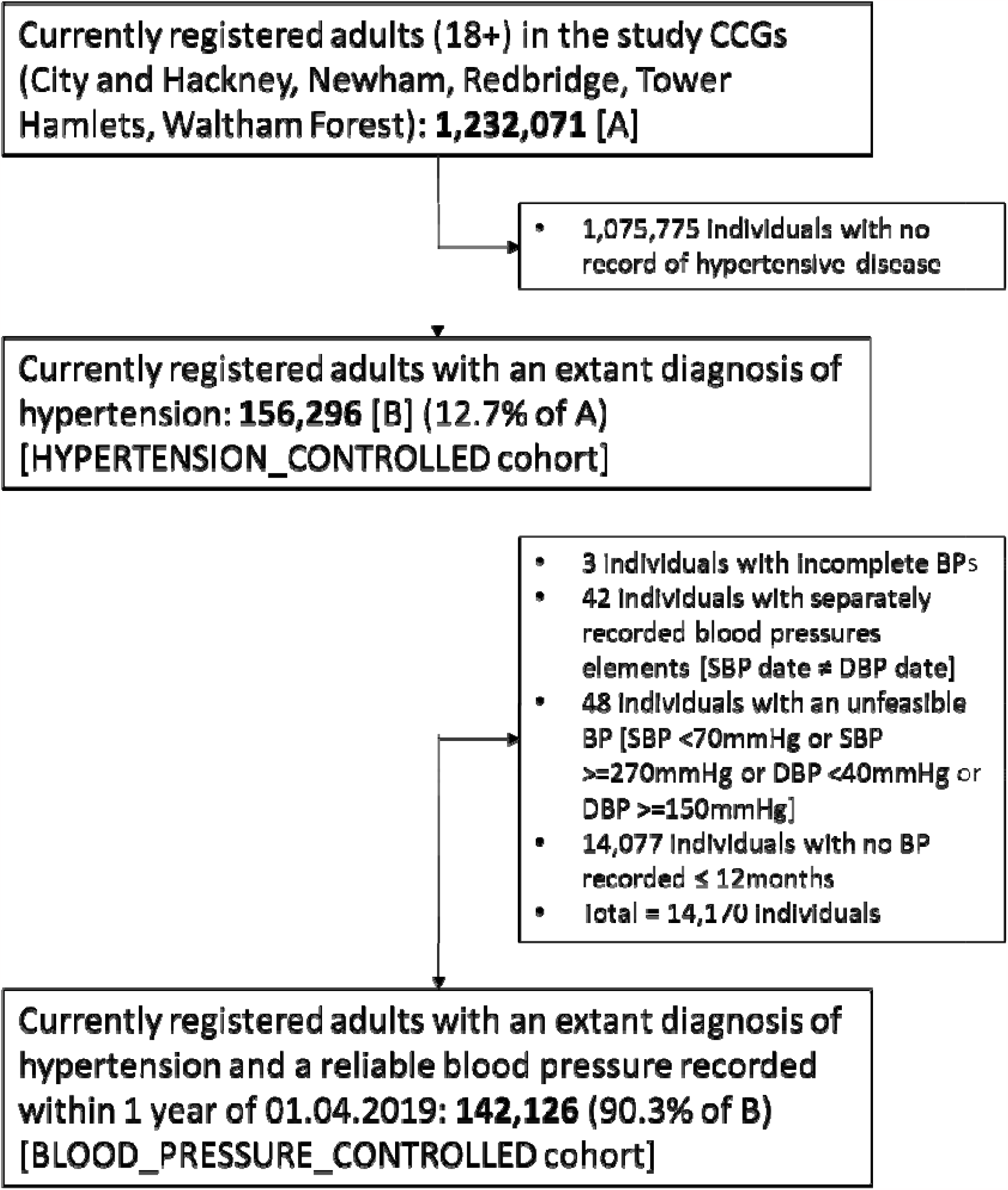
Study population flowchart.

Medicines prescribed with 6 months of the index date were considered for eight classes of antihypertensive medication (Supplementary Table 2): i) ACE inhibitors/Angiotensin Receptor Blockers, ii) Beta-blockers, iii) Potassium-sparing diuretics, iv) Calcium channel blockers, v) Thiazide-type and thiazide-like diuretics, vi) Centrally-acting anti-hypertensives, vii) Alpha-blockers and viii) Loop diuretics. The number of different classes prescribed were grouped into categoric treatment intensity: 0, 1, or 2 or more medications.

Ethnic groups were categorised according to Office of National Statistics 2001 census categories and comprised White – including White British, Irish, or White other; Black including Black British, Caribbean, African, and other Black background; Asian including British Asian, Bangladeshi, Pakistani, Indian and any other Asian background; Chinese and other ethnic groups were classified as Other ethnic groups. Mixed ethnic groups were categorised as Mixed and Unknown comprised no ethnicity code recorded, unclassified codes or not stated.[15]

Index of Multiple Deprivation was based on 2019 small area indicators (LSOA) and used the national quintiles from quintile 1 (most deprived) to quintile 5 (least deprived).[16]

Blood pressure was deemed controlled as per NHS QOF indicators HYP003 and HYP007: individuals under 80 years of age: systolic blood pressure less than or equal to 140mmHg and diastolic blood pressure less than or equal to 90mmHg (HYP003); individuals 80 years of age and older, systolic blood pressure less than or equal to 150mmHg and diastolic blood pressure less than or equal to 90mmHg (HYP007).[17]

## Outcomes

Three binary outcome variables of blood pressure management were considered.

1. BLOOD_PRESSURE_RECORDED was true of Individuals with a valid blood pressure recorded within 12 months of the index date.
2. BLOOD_PRESSURE_CONTROLLED considered only Individuals with a blood pressure recorded within 12 months of the index date and was true for patients with a blood pressure within the QOF age-adjusted target.
3. HYPERTENSION_CONTROLLED considered the entire cohort and was true for patients with a blood pressure within the QOF age-adjusted target. Individuals without a blood pressure within 12 months of the index date were considered to be above target blood pressure.

## Statistics

Data were processed, aggregated and validated and descriptive statistics derived using Python (version 3.9.1). Univariate and multiple logistic regressions analyses (adjusted for Sex, Age, Ethnicity, IMD quintile and treatment intensity) were performed using R (version 4.0.5) with sub-analyses performed by treatment intensity. Forest plots were generated using the forestplot python package (version 0.2.0).[18]

### Information governance and ethics

This was confirmed by the Chair of the North East London Strategic Information Governance Network. The clinical effectiveness group (CEG) has the written consent of all practices in the study area to use pseudonymised patient data for audit and research for patient benefit. The CEG is the data processor, and the General Practices in the study are the data controllers. The researchers adhere to the data protection principles of the Data Protection Act 2018, and all data was managed according to UK NHS information governance requirements. All data were pseudonymised and are only presented in aggregate form. The NHS Health Research Authority toolkit (http://www.hra-decisiontools.org.uk/ethics/) identified that Research Ethics Approval was not required for this project. This was confirmed by the Chair of the North East London Strategic Information Governance Network.

## RESULTS

The study cohort was derived from 190 out of 199 practices in the participating five North-East London localities with a population of approximately 1.23 million adults of whom 156,296 had hypertension. Nine practices were excluded as they used a different electronic health record system not available to the researchers.

The summary characteristics are described in Table 1. Among the whole cohort, 90.0% (142,126/156,296) had a valid blood pressure recorded in the 12 months prior to the index date (BLOOD_PRESSURE_RECORDED). 14,077 individuals did not have a blood pressure recorded within 12 months of the index date and 93 blood pressure recordings were missing (Figure 1).

**Table 1:**
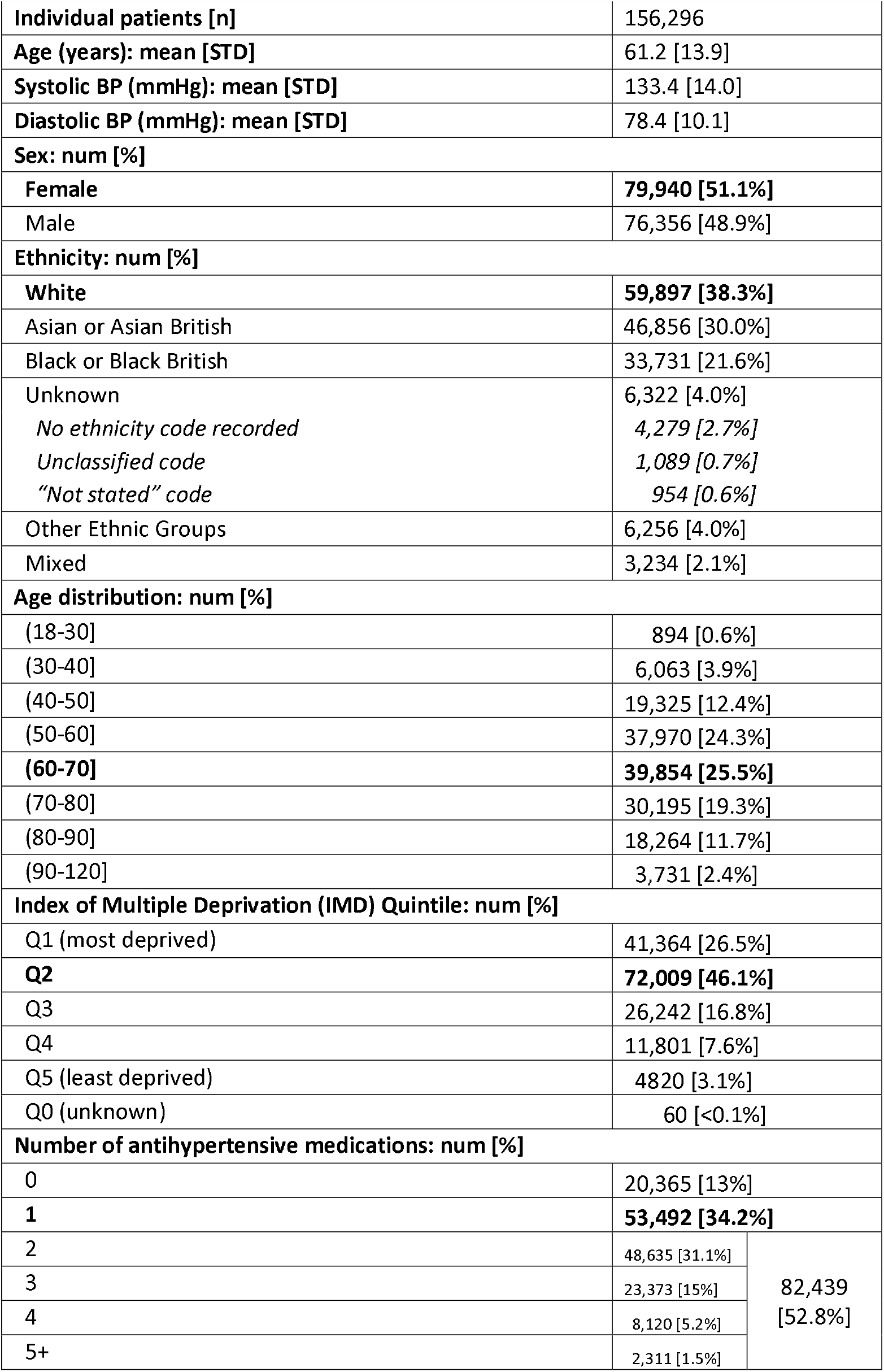
Characteristics of patients included in the study cohort (N= 156,296 adult patients with hypertension). In variables with multiple categories, the largest top-level category is shown in bold type.

Of those with recorded blood pressure, 80.7% (114,653/142,126) had controlled BP as per age-adjusted targets (BLOOD_PRESSURE_CONTROLLED). Using the entire hypertension register (assuming all missing values were uncontrolled), HYPERTENSION_CONTROLLED was 73.4% (114,653/156,296) controlled to age-adjusted targets.

### Blood pressure recording and ethnicity

BLOOD_PRESSURE_RECORDED within 12 months and ethnicity were modelled using unadjusted and adjusted logistic regression in relation to the White ethnic groups. Asian ethnic groups were 60% more likely to have a recent blood pressure record than the White ethnic group (Odds Ratio (OR) 1.60, 95% CI = 1.53-1.68). Other ethnic groups (except for the Unknown ethnicity group) were all within 10% of the reference White ethnic group (Table 2A and Supplementary Figure 1A).

**Table 2:**
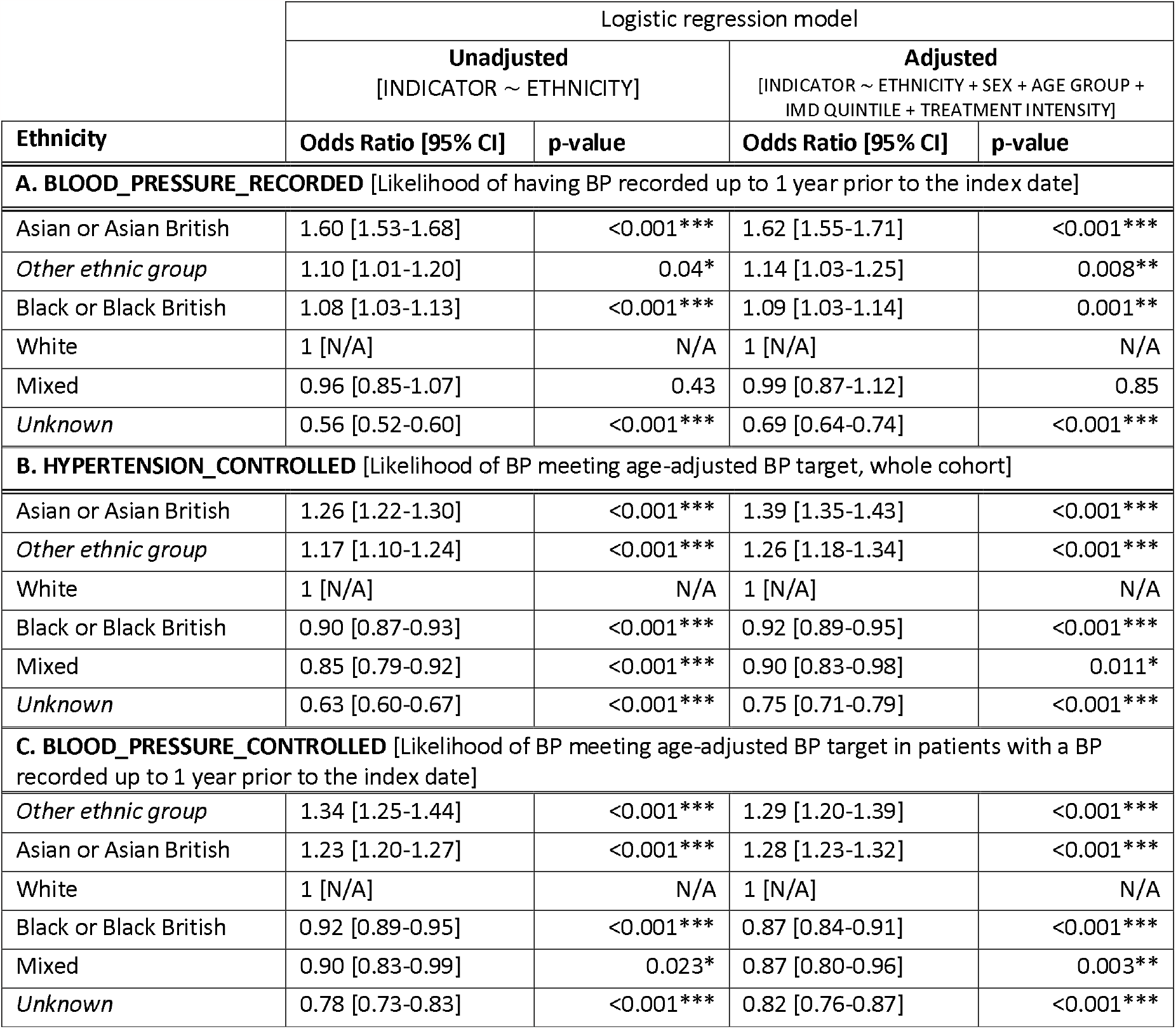
Binomial logistic regression modelling of the impact of ethnicity on blood pressure control as assessed by the A. BLOOD_PRESSURE_RECORDED, B. HYPERTENSION_CONTROLLED, and C. BLOOD_PRESSURE_CONTROLLED indicators. (* p-val <0.05, ** p-val <0.01, *** p-val <0.001).

In the multivariate model, the odds ratios for blood pressure recording were essentially unchanged. For the Asian ethnic groups, the OR was 1.62 (95% CI = 1.55 to 1.71), and for Black ethnic groups, the OR was 1.09, 95% CI = 1.03-1.14) as shown in Figure 2A.

**Figure 2:**
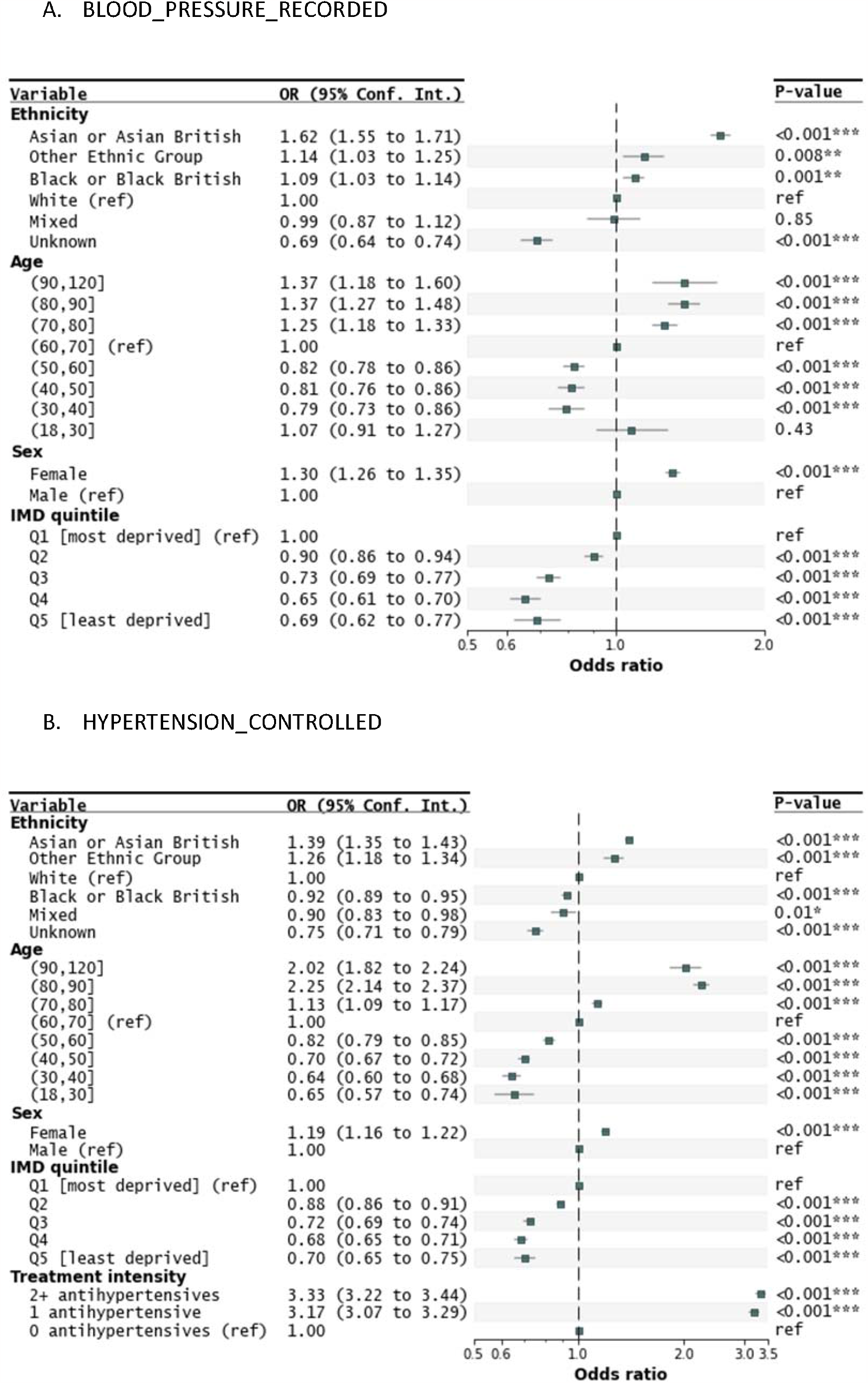
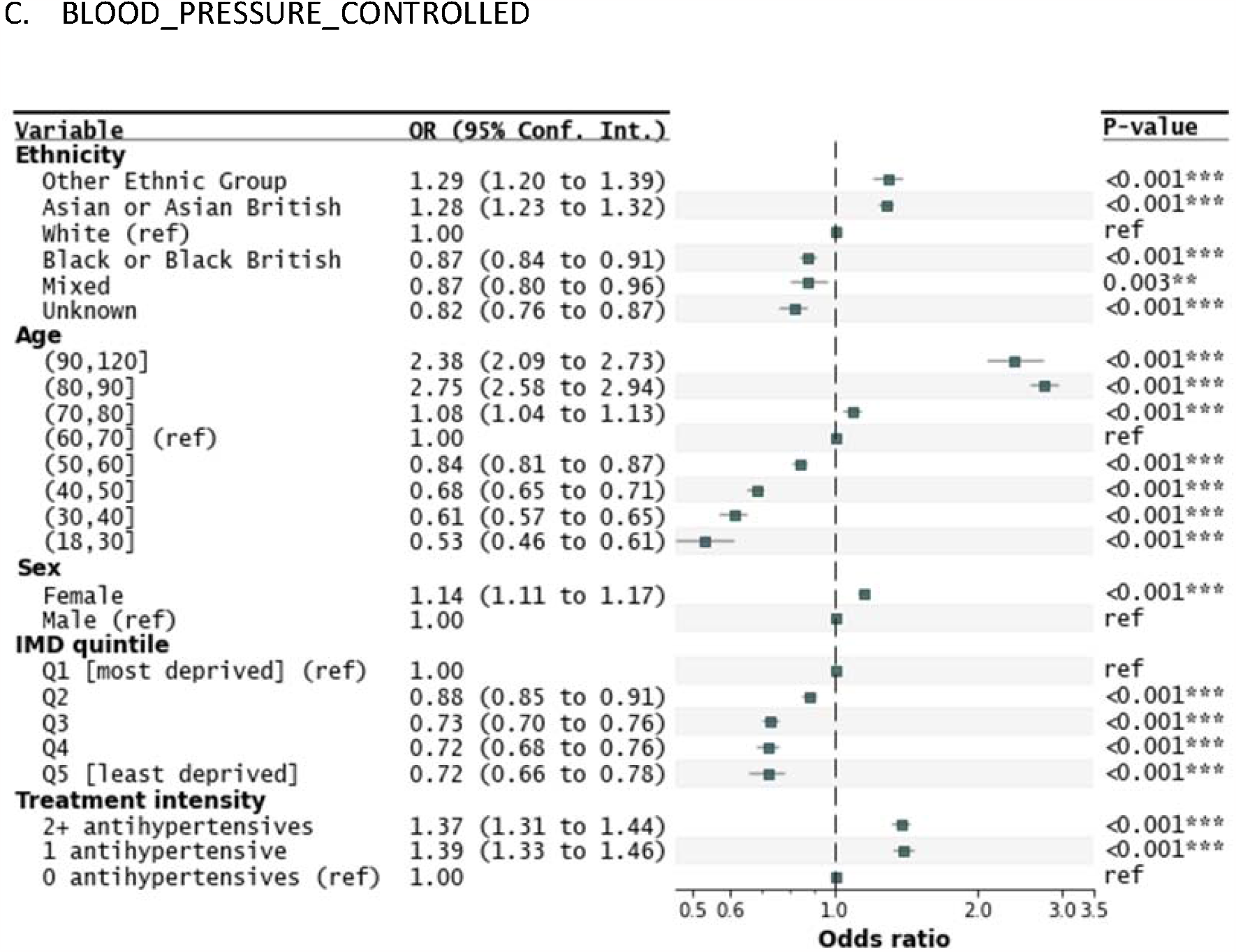
Forest plots of Odds Ratios (ORs) for A. BLOOD_PRESSURE_RECORDED, B. HYPERTENSION_CONTROLLED and C. BLOOD_PRESSURE_CONTROLLED. The squares plot the OR relative to the reference (ref) category and the whisker bars are the 95% confidence interval for the OR. Squares to the left of the vertical line (OR = 1.0) indicate categories in which individuals are less likely to have a blood pressure recorded within 12 months or the index date (A) or controlled blood pressure (B and C), whilst squares on the right indicate individuals more likely to have a recorded blood pressure (A) or a controlled blood pressure (B and C). HYPERTENSION_CONTROLLED (B) considers the whole cohort whilst BLOOD_PRESSURE_CONTROLLED (C) considers only cohort members with a valid blood pressure recorded within 12 months of the index date. The ORs for treatment intensity for BLOOD_PRESSURE_RECORDED are not plotted as they are significantly larger than the other ORs (1 medication: 7.83; 2+ medications: 9.99) and affect graph clarity.

### Control of hypertension and ethnicity

HYPERTENSION_CONTROLLED (where individuals with no blood pressure recorded within 12 of the index date were categorised as uncontrolled blood pressure) and ethnicity were likewise modelled using univariate and multivariate models (Table 2B).

In the univariate model, the Asian ethnic groups were 26% more likely to have controlled hypertension than the White ethnicity group (OR 1.26, 95% CI = 1.22 to 1.30) and the Black ethnic groups were 10% less likely (OR 0.90, 95%CI = 0.87 to 0.93) to have controlled hypertension than the White ethnic groups (Supplementary Figure 1B).

In the multivariate model (Figure 2B), the Asian ethnic groups were 39% more likely to have controlled hypertension than the White ethnic groups (OR 1.19, 95% = CI 1.35-1.43) whilst the Black ethnic groups were 8% less likely (OR 0.92, 95% CI 0.89-0.95).

### Control of blood pressure and ethnicity

The relationship between BLOOD_PRESSURE_CONTROLLED was modelled as for the two previous indicators (Table 2**Error! Reference source not found**.C).

In the univariate model, the Asian ethnic group were 23% more likely to have controlled blood pressure than the White ethnic group (OR 1.23, 95% CI = 1.20 to 1.27) whereas the Black ethnic groups were 8% less likely (OR 0.92, 95% CI = 0.89 to 0.95) as shown in Supplementary Figure 1C. These differences were further increased in the multivariate model (Figure 2C). The Asian ethnic groups were 28% more likely to have controlled blood pressure than White ethnic groups (OR 1.28, 95% CI = 1.23 to 1.32); the Black ethnic groups were 13% less likely to have a controlled blood (OR 0.87, 95% CI = 0.84 to 0.91).

### Sex, age, treatment intensity and IMD

The likelihood of controlled blood pressure increased with age (Figure 2C). Compared to individuals in the (most populated) 60-70 years age-bracket (25.5% [39,854/156,296]), individuals in the 30-40 years age bracket (3.9% [6,063/156,296]) were 39% less likely to have a controlled blood pressure (OR 0.61, 95% CI = 1.57 to 1.65). Conversely, individuals in the 80-90 years bracket (11.7% [18,264/156,296]) were 175% (OR 2.75, 95%CI = 2.58 to 2.94) more likely to have controlled blood pressure. Patients under 50 years of age were 40% less likely to have controlled blood pressure than those 50 years and older (OR 0.60, 95% CI =0.60-0.61, p-value = <0.001).

The likelihood of blood pressure control decreased with decreasing deprivation (Figure 2C). Patients in the least deprived Index of Multiple Deprivation quintiles (Q4 and Q5) were 28% less likely than patients in the most deprived quintile (Q1) to have controlled blood pressure (ORs 0.72, CI = 0.68 to 0.76 and 0.72, 95% CI = 0.66 to 0.78 respectively). Women were 14% more likely to have controlled blood pressure than men (Figure 2C; OR 1.14. 95% CI = 1.11 to 1.17).

Relative to untreated individuals, the likelihood of blood pressure control increased with treatment intensity by approximately 38% regardless of treatment intensity (ORs: 1 antihypertensive 1.39, 95% CI = 1.33 to 1.46; 2+ antihypertensives 1.37, 95% CI = 1.31 to 1.44, Figure 2C).

### Ethnicity and treatment intensity

The distribution of treatment intensities by ethnic group in those with controlled and uncontrolled recorded blood pressure is shown in Supplementary Figure 2.

Asian ethnic groups were the least likely to be untreated in both the uncontrolled and controlled blood pressure groups: 10.6% (908/7,639) and 7.3% (2,627/36,169) respectively. Black ethnic groups were similarly or less likely to be untreated be on a single anti-hypertensive medication than the White ethnic groups. In the controlled blood pressure group, 8.5% (2,016/23,850) of the Black ethnic groups were untreated and 32.9% (7,851/23,850) on a single agent (cf. 9.4% (4,084/43,659) and 34.4% (15,033/43,659) for the White ethnic groups). In the uncontrolled blood pressure group, 13.6% (918/6,731) of the Black ethnic groups were untreated and 31.7% (2,133/6,731) on a single agent as compared to 13.0% (1,329/10,233) and 36.0% (3,685/10,233) for the White ethnic groups.

A multivariate analysis was conducted for the same indicator (BLOOD_PRESSURE_CONTROLLED) was repeated on: i) untreated individuals, ii) individuals on one class of antihypertensive, and iii) individuals on two or more classes of antihypertensives. The ORs by ethnic group for this sub-analysis are shown in Figure 3. Relative to White ethnic groups, and regardless of the number of medications they were on, the Asian ethnic groups were more 12-26% more likely to have controlled blood pressure: 0 medications: OR 1.26, 95% CI = 1.13 to 1.40; 1 medication: OR 1.18, 95% CI = 1.12 to 1.25; 2+ medications: OR 1.22, 95% CI = 1.16 to 1.28. Black ethnicity groups were always 10-23% less likely to have controlled blood pressure (0 medications: OR 0.77, 95% CI = 0.69 to 0.85; 1 medication: OR 0.90, 95% CI = 0.85 to 0.96; 2+ medications: OR 0.85, 95% CI = 0.81 to 0.90.

**Figure 3:**
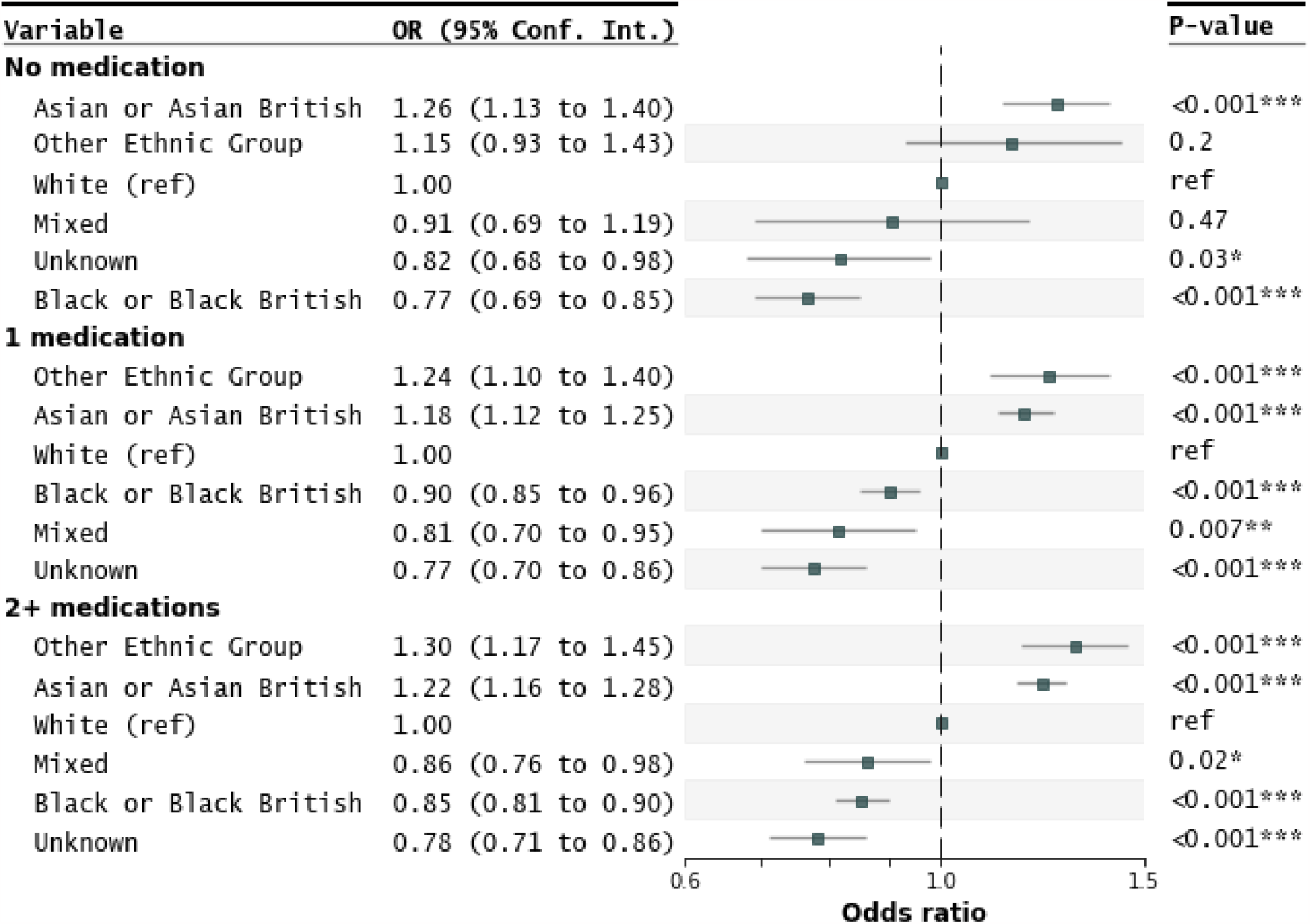
Odds ratios (BLOOD_PRESSURE_CONTROLLED) by ethnicity for patients on 0, 1, and 2 or more (2+) medications.

## DISCUSSION

### Summary

This cross-sectional study of an unselected, large cohort of 156,296 adult patients with hypertension from 190 GP practices in North-East London, identified inequities in treatment of hypertension by ethnicity, age, sex and deprivation.

Black ethnic groups were more likely to have uncontrolled hypertension than White or Asian ethnic groups. This was not due to less frequent recording of blood pressure in Black groups who were in fact more likely to have a blood pressure recorded within 1 year than White ethnic groups, nor was this finding due to associations with age, sex or deprivation. Regardless of treatment intensity, control of blood pressure in the Black ethnic groups was worse and in Asian ethnic groups better, than in White ethnic groups.

These data suggest that lower levels of blood pressure control in the Black ethnic groups cannot be directly linked to relative under-recording of blood pressure or less intensive treatment. Possible other possible factors include unidentified confounding factors, differences in age at diagnosis and treatment escalation, and physiological treatment resistance. [10,19–21] Medication costs on working age adults and adherence may also be contributory factors in particularly in younger people.[22,23]

Patients with hypertension under 50 years were 40% less likely than those older, to have controlled hypertension. Given that 17% of the cohort was under the age of 50 with more potential years at risk, this represents a notable treatment inequity.[22] Similar concerns have been expressed about statin under-treatment.[24]

Lastly, this study highlights the importance of indicator choice in understanding control of hypertension. The difference between BLOOD_PRESSURE_CONTROLLED and the QOF derived HYPERTENSION_CONTROLLED indicators has been previously noted as reflecting missing recordings rather than reduction in control. [4] QOF indicators may be suitable for performance purposes, but do not provide satisfactory indicators of population blood pressure control.

### Strengths and limitations

The study cohort is unselected, including almost all (95%) general practices in the region with high levels (96%) of recorded self-reported ethnicity in an ethnically diverse population broad representation of the three principal ethnic groups discussed. However, it is important to me mindful that high level ethnic groupings can conceal underlying heterogeneity between constituent ethnic groups.[25]

The study was cross-sectional and trajectories of the patient groups through time were not addressed. Treatment changes within 6 months would not be captured accurately, possibly overestimating treatment intensity in a small number of patients. In North East London 72.6% of the population were in the two most deprived quintiles and only 10% in the two most affluent. The urban density and the fact that the rich and poor often live in close proximity means that area level measures such as IMD quintile reduce gradients between extremes of deprivation. In addition, the most affluent may be away from their London residences for prolonged periods or access alternative health services which may further influence blood pressure recording and control and contribute to apparent poor management in this group.

### Comparison with existing literature

Our study confirms previous findings of inequities in the control of blood pressure in patients with hypertension, with poorer control in Black ethnic groups, and better control in Asian ethnic groups in comparison to White groups as well as better control in women compared to men.[8–12] We found younger groups had poorer control than older groups, a finding not replicated in Biobank though that study did not consider individuals under 40 years of age.[9] In our study, Black ethnic groups had similar access to primary-care based blood pressure recording as other ethnic groups, a finding similar to that in South East London.[8]

In a UK-wide study by Eastwood *et al*. comparing management of newly diagnosed hypertension in European, South Asian and African/African Caribbean ethnicities, the findings of poorer control in Black ethnic groups was similar to our own study but the South Asian group were found to have similar levels of BP monitoring and control in comparison to other ethnic groups.[10] We found higher levels of BP monitoring and of BP control in Asian than White ethnic groups. However, the UK-wide study considered only recently diagnosed patients which may account for differences in findings in South Asian groups.

Our study showed that, regardless of treatment intensity, Black ethnic groups were less likely to have controlled hypertension. Eastwood *et al*. did not find differences between BP control in the African/African Caribbean and European groups when considering only antihypertensive-adherent people suggesting that adherence might be a factor, with cost of prescriptions as a possible contributing factor in working adults.[26]

### Implications for research and practice

In North-East London and the UK more generally, a focus on the optimisation of blood pressure control in Black ethnic groups and younger people would be an important step in addressing outcome inequity. The impact of medication costs in younger working age-groups deserves further attention and in particular the role of single pill combination therapies that reduce patient costs and improve adherence. Easier access to monitoring of blood pressure is relevant to all people with hypertension particularly those of working age. [27–29]

## Supporting information

RECORD Checklist

## Data Availability

All data produced in the present study are available upon reasonable request to the authors

## ETHICS APPPOVAL

Based on the NHS Health Research Authority Questionnaire (http://www.hra-decisiontools.org.uk/ethics/) Research Ethics approval was not required for this project as patient level data are anonymised, and only aggregated patient data are reported in this study. All GPs in the participating east London practices consented to the use of their anonymised patient data for research and development for patient benefit. This was confirmed by the Chair of the North East London Strategic Information Governance Network.

## COMPETING INTERESTS

The authors have declared no competing interests.

## ACKNOWLEDGMENTS

We are grateful to the general practitioners and their practice teams for allowing the use of their patient records, to the Clinical Effectiveness Group for providing access to their curated high-quality dataset and to the population in East London from whom the data are derived. The authors wish to thank staff at CEG for supporting practices with guidance and data entry tools which support this project.

We are grateful for Lucas Shen’s help with his forestplot Python package.

## FUNDING

This work was supported by Barts Charity and Health Data Research UK, an initiative funded by UK Research and Innovation, Department of Health and Social Care (England) and the devolved administrations, and leading medical research charities. OR is supported by the National Institute for Health Research (NIHR) and a Drayson research fellowship. RM is supported by Barts Charity (MGU0504). ZR-E is supported by an NIHR Integrated Academic Training programme and her Academic Clinical Lectureship post and is also supported by a British Heart Foundation Clinical Research Training Fellowship (FS/17/81/33318). CC is supported by an NIHR School of Primary Care Research Clinical Fellowship Queen Mary University of London. The views expressed are those of the authors and not necessarily those of the NIHR or the Department of Health and Social Care.

## DATA SHARING

All data relevant to the study are included in the article.

## AUTHOR CONTRIBUTIONS

The study was designed by SR and JR. Data analysis was by SR, ID and OR. The report was written by SR and JR with contributions from all authors.

