## Supplementary material for "Inequities in hypertension management: an observational cross-sectional study in North-East London using electronic health records"

**BJGP**

SUPPLEMENTARY MATERIAL

**Authors:**

- Stuart Rison^1,2^
- Oliver Redfern^3^
- Isabel Dostal^1^
- Chris Carvalho^1,2^
- Rohini Mathur^1^
- Zahra Raisi-Estabragh^4,5^
- John Robson^1^

**Affiliations:**

1. Centre for Primary Care, Wolfson Institute of Population Health, Barts and The London School of Medicine and Dentistry, Queen Mary University of London, London, UK.
2. North East London Integrated Care System, Unex Tower, London, UK.
3. Nuffield Department of Clinical Neurosciences, University of Oxford, Oxford, UK
4. Barts Heart Centre, St Bartholomew's Hospital, Barts Health NHS Trust, London, UK
5. William Harvey Research Institute, National Institute for Health and Care Research Barts Biomedical Research Centre, Queen Mary University London, London, UK.

### Supplementary tables

| Read v2 reference for selected codes in the Readv2 definition | | List of included SNOMED CT codes equivalent to selected codes in the Readv2 definition | | |
| --- | --- | --- | --- | --- |
| **Read code** | **Read term** | **SNOMED CT code** | **SNOMED CT concept ID** | **SNOMED CT description ID** |
| G2 | Hypertensive disease | Hypertensive disease | 38341003 | 64168014 |
|  |  | High blood pressure | 38341003 | 64172013 |
|  |  | Hypertensive vascular disease | 38341003 | 64173015 |
|  |  | Hypertensive vascular degeneration | 38341003 | 64174014 |
|  |  | BP - High blood pressure | 38341003 | 490277011 |
|  |  | Systemic arterial hypertension | 38341003 | 490278018 |
|  |  | HBP - High blood pressure | 38341003 | 490280012 |
|  |  | HT - Hypertension | 38341003 | 490281011 |
|  |  | High blood pressure disorder | 38341003 | 490282016 |
|  |  | BP+ - Hypertension | 38341003 | 490283014 |
|  |  | HTN - Hypertension | 38341003 | 2164904016 |
|  |  | Hypertensive disorder, systemic arterial | 38341003 | 2671386015 |
|  |  | BP - hypertensive disease | 38341003 | 523801000006119 |
| G20 | Essential hypertension | Essential hypertension | 59621000 | 99042012 |
|  |  | Idiopathic hypertension | 59621000 | 99044013 |
|  |  | Systemic primary arterial hypertension | 59621000 | 99046010 |
|  |  | Benign essential hypertension | 1201005 | 3135013 |
|  |  | Diastolic hypertension | 48146000 | 80224019 |
|  |  | Systolic hypertension | 56218007 | 93494011 |
|  |  | Malignant essential hypertension | 78975002 | 131046010 |
|  |  | Accelerated essential hypertension | 78975002 | 503982017 |
|  |  | Malignant hypertension | 78975002 | 884121000006111 |
|  |  | Essential hypertension NOS | 59621000 | 648911000006113 |
|  |  | Hypertension | 38341003 | 64176011 |
|  |  | High blood pressure | 38341003 | 64172013 |
|  |  | Hypertensive vascular disease | 38341003 | 64173015 |
|  |  | Hypertensive vascular degeneration | 38341003 | 64174014 |
|  |  | BP - High blood pressure | 38341003 | 490277011 |
|  |  | Systemic arterial hypertension | 38341003 | 490278018 |
|  |  | HBP - High blood pressure | 38341003 | 490280012 |
|  |  | HT - Hypertension | 38341003 | 490281011 |
|  |  | High blood pressure disorder | 38341003 | 490282016 |
|  |  | BP+ - Hypertension | 38341003 | 490283014 |
|  |  | HTN - Hypertension | 38341003 | 2164904016 |
|  |  | Hypertensive disorder, systemic arterial | 38341003 | 2671386015 |
|  |  | Elevated blood pressure | 24184005 | 196353013 |
|  |  | Finding of increased blood pressure | 24184005 | 196354019 |
|  |  | Blood pressure elevation | 24184005 | 2920698012 |
|  |  | Primary hypertension | 59621000 | 99047018 |
| G24 | Secondary hypertension | Secondary hypertension | 31992008 | 53452019 |
|  |  | Secondary benign hypertension | 194785008 | 299678018 |
|  |  | Benign secondary hypertension | 194785008 | 2470031013 |
|  |  | Secondary benign renovascular hypertension | 73410007 | 2478822013 |
|  |  | Benign secondary renovascular hypertension | 73410007 | 121910014 |
|  |  | Hypertension secondary to endocrine disorders | 194788005 | 299681011 |
|  |  | Hypertension secondary to endocrine disorder | 194788005 | 2532161014 |
|  |  | Endocrine hypertension | 194788005 | 3763241013 |
|  |  | Malignant secondary hypertension | 89242004 | 147988014 |
|  |  | Accelerated secondary hypertension | 89242004 | 508416019 |
|  |  | Secondary malignant renovascular hypertension | 194783001 | 299676019 |
|  |  | Malignant secondary renovascular hypertension | 194783001 | 2470030014 |
|  |  | Secondary malignant hypertension NOS | 89242004 | 151171000006110 |
| G2y | Other specified hypertensive disease | Other specified hypertensive disease | 38341003 | 10441000006116 |
|  |  | High blood pressure | 38341003 | 64172013 |
|  |  | Hypertensive vascular disease | 38341003 | 64173015 |
|  |  | Hypertensive vascular degeneration | 38341003 | 64174014 |
|  |  | BP - High blood pressure | 38341003 | 490277011 |
|  |  | Systemic arterial hypertension | 38341003 | 490278018 |
|  |  | HBP - High blood pressure | 38341003 | 490280012 |
|  |  | HT - Hypertension | 38341003 | 490281011 |
|  |  | High blood pressure disorder | 38341003 | 490282016 |
|  |  | BP+ - Hypertension | 38341003 | 490283014 |
|  |  | HTN - Hypertension | 38341003 | 2164904016 |
|  |  | Hypertensive disorder, systemic arterial | 38341003 | 2671386015 |
|  |  | Stage 1 hypertension | 1806071000006102 | 1806071000006118 |
|  |  | Stage 2 hypertension | 1806081000006104 | 1806081000006115 |
|  |  | Severe hypertension | 1806141000006109 | 1806141000006113 |
| G2z | Hypertensive disease NOS | Hypertensive disorder | 38341003 | 1215744012 |
|  |  | High blood pressure | 38341003 | 64172013 |
|  |  | Hypertensive vascular disease | 38341003 | 64173015 |
|  |  | Hypertensive vascular degeneration | 38341003 | 64174014 |
|  |  | BP - High blood pressure | 38341003 | 490277011 |
|  |  | Systemic arterial hypertension | 38341003 | 490278018 |
|  |  | HBP - High blood pressure | 38341003 | 490280012 |
|  |  | HT - Hypertension | 38341003 | 490281011 |
|  |  | High blood pressure disorder | 38341003 | 490282016 |
|  |  | BP+ - Hypertension | 38341003 | 490283014 |
|  |  | HTN - Hypertension | 38341003 | 2164904016 |
|  |  | Hypertensive disorder, systemic arterial | 38341003 | 2671386015 |
| Gyu2 | [X]Hypertensive diseases | [X]Hypertensive diseases | 38341003 | 389341000006117 |
|  |  | High blood pressure | 38341003 | 64172013 |
|  |  | Hypertensive vascular disease | 38341003 | 64173015 |
|  |  | Hypertensive vascular degeneration | 38341003 | 64174014 |
|  |  | BP - High blood pressure | 38341003 | 490277011 |
|  |  | Systemic arterial hypertension | 38341003 | 490278018 |
|  |  | HBP - High blood pressure | 38341003 | 490280012 |
|  |  | HT - Hypertension | 38341003 | 490281011 |
|  |  | High blood pressure disorder | 38341003 | 490282016 |
|  |  | BP+ - Hypertension | 38341003 | 490283014 |
|  |  | HTN - Hypertension | 38341003 | 2164904016 |
|  |  | Hypertensive disorder, systemic arterial | 38341003 | 2671386015 |
|  |  | [X]Other secondary hypertension | 31992008 | 413461000006118 |
|  |  | [X]Hypertension secondary to other renal disorders | 31992008 | 389331000006110 |
| G25 | Stage 1 hypertension (NICE - Nat Ins for Hth Clin Excl 2011) | Stage 1 hypertension (NICE - National Institute for Health and Clinical Excellence 2011) | 843821000000102 | 2189411000000111 |
|  |  | Stage 1 hypertension (NICE 2011) without evidence of end organ damage | 908631000000108 | 2335761000000116 |
|  |  | Stage 1 hypertension (NICE 2011) with evidence of end organ damage | 908651000000101 | 2335801000000114 |
|  |  | Stage 1 hypertension | 843821000000102 | 2193031000000112 |
| G26 | Severe hypertension (Nat Inst for Health Clinical Ex 2011) | Severe hypertension (NICE - National Institute for Health and Clinical Excellence 2011) | 843841000000109 | 2189451000000110 |
|  |  | Severe hypertension | 843841000000109 | 2193021000000110 |
| G28 | Stage 2 hypertension (NICE - Nat Ins for Hth Clin Excl 2011) | Stage 2 hypertension (NICE - National Institute for Health and Clinical Excellence 2011) | 846371000000103 | 2194941000000119 |
|  |  | Stage 2 hypertension | 846371000000103 | 2211211000000110 |
| *Hypertension resolved codes* | | | | |
| 21261 | Hypertension resolved | Hypertension resolved | 162659009 | 253532015 |
| 212K | Hypertension resolved | Hypertension resolved | 162659009 | 975021000006111 |

Table S 1: Hypertension codes (deposited on ClinicalCodes.org on 03/02/2023: https://clinicalcodes.rss.mhs.man.ac.uk/medcodes/article/200/).

| **Datum** | **Category** |
| --- | --- |
| Age | Patient Details |
| Gender | Patient Details |
| Lower Layer Area (2011) | Patient Details |
| Organisation Code | Patient Details |
| Ethnicity | Clinical Code |
| Hypertension | Clinical Code |
| Systolic BP | Clinical Code |
| Systolic BP | Date |
| Systolic BP | Value |
| Diastolic BP | Clinical Code |
| Diastolic BP | Date |
| Diastolic BP | Value |
| ACEi/ARB | Name, Dosage and Quantity |
| ACEi/ARB | Date of Issue |
| Beta Blocker | Name, Dosage and Quantity |
| Beta Blocker | Date of Issue |
| K-Sparing | Name, Dosage and Quantity |
| K-Sparing | Date of Issue |
| CCBs | Name, Dosage and Quantity |
| CCBs | Date of Issue |
| Thiazide Diuretic | Name, Dosage and Quantity |
| Thiazide Diuretic | Date of Issue |
| Centrally Acting HTs | Name, Dosage and Quantity |
| Centrally Acting HTs | Date of Issue |
| Alpha Blockers | Name, Dosage and Quantity |
| Alpha Blockers | Date of Issue |
| Loop Diuretic | Name, Dosage and Quantity |
| Loop Diuretic | Date of Issue |

Table S 2: Patient data collected.

### Supplementary figures

A. **HYPERTENSION_CONTROLLED**

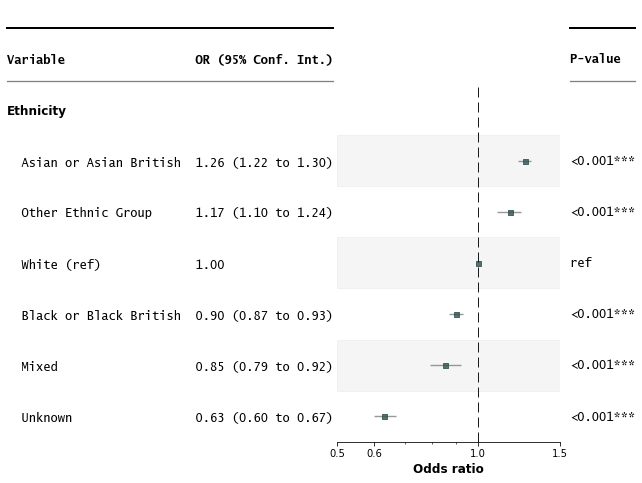

B. **BLOOD_PRESSURE_RECORDED**

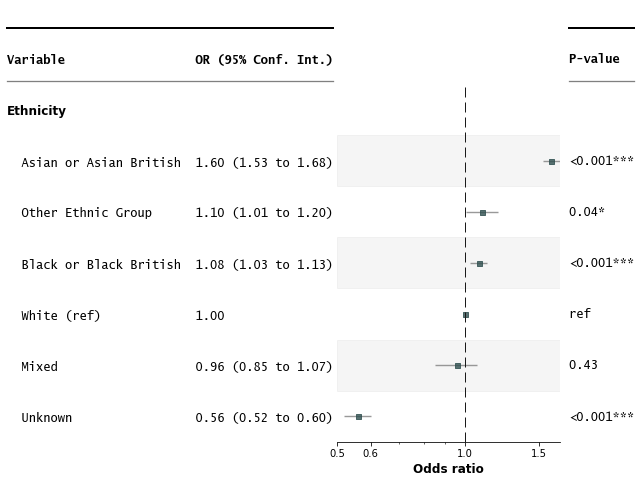

C. **BLOOD_PRESSURE_CONTROLLED**

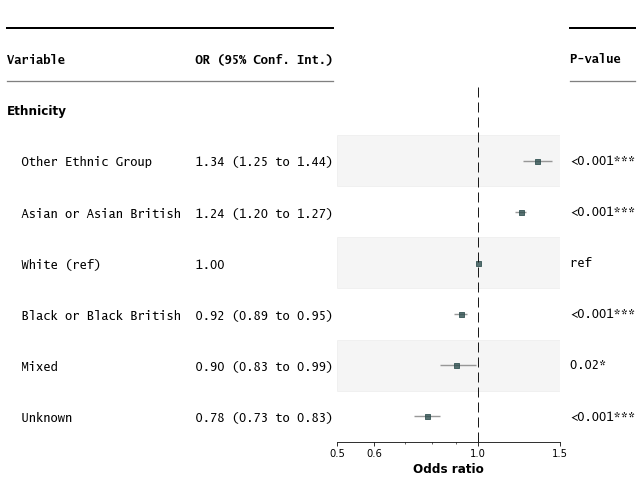

Figure S 1: Odd ratios forest plots for univariate analyses (INDICATOR ~ ETHNICITY) for the A. HYPERTENSION_CONTROLLED, B. BLOOD_PRESSURE_RECORDED and C. BLOOD_PRESSURE_CONTROLLED.

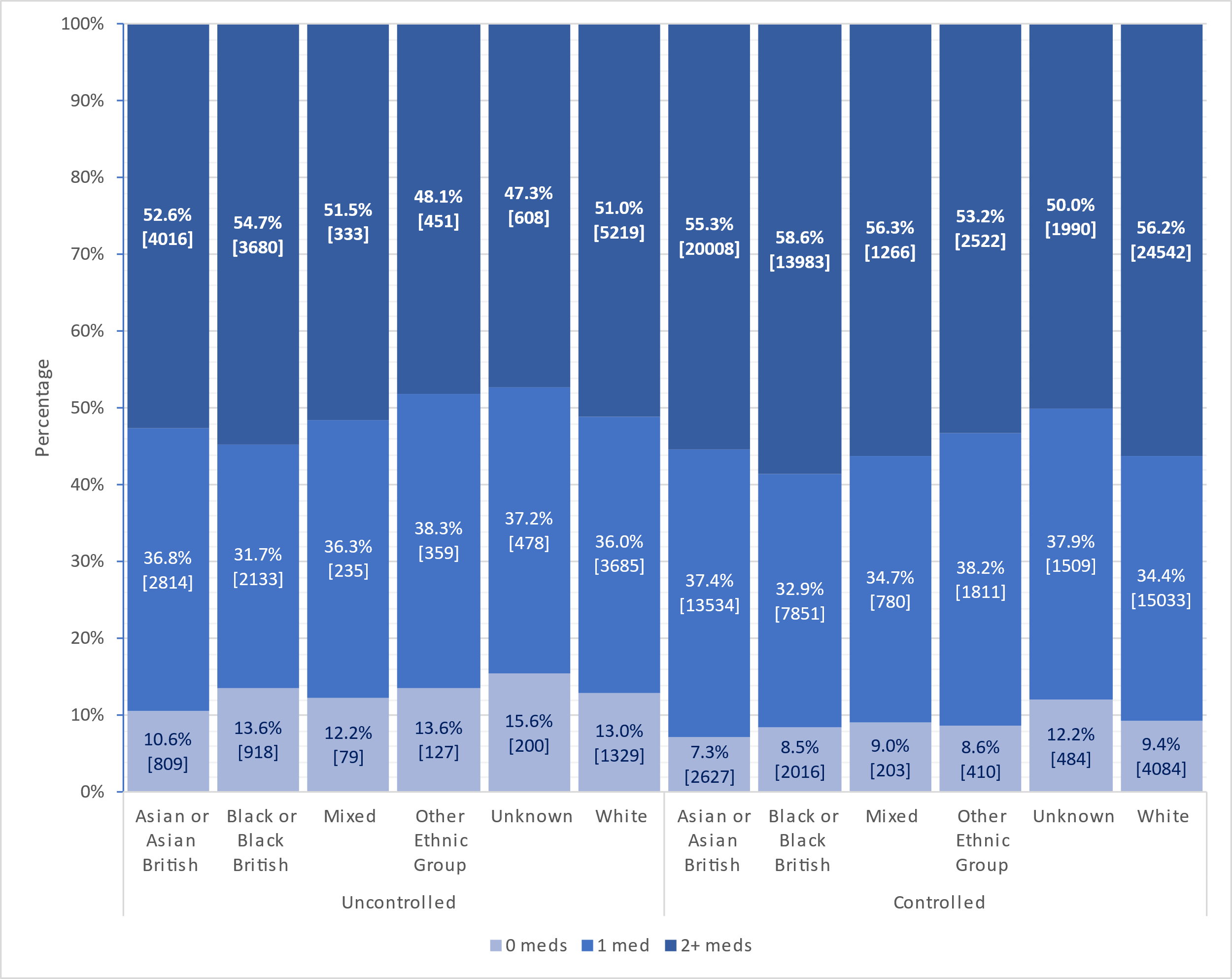

Figure S 2 Treatment intensities by ethnic groups. The percentage of patient with no (0 meds), 1 (1 med) or two or more (2+ meds) classes of antihypertensives prescribed within 6 months of the index date (1st April 2019). The 6 bars on the left show distribution of treatment intensity in patients with controlled blood pressure, the 6 bars on the right, for patients with uncontrolled blood pressure. Actual number of patients are given below the percentages.
